# Molecular epidemiological characteristics of *Mycobacterium abscessus* complex in non-cystic fibrosis patients in Japan and Taiwan

**DOI:** 10.1101/2021.10.23.21265405

**Authors:** Mitsunori Yoshida, Jung-Yien Chien, Kozo Morimoto, Takeshi Kinjo, Akio Aono, Yoshiro Murase, Keiji Fujiwara, Yuta Morishige, Hiroaki Nagano, Ruwen Jou, Naoki Hasegawa, Manabu Ato, Yoshihiko Hoshino, Po-Ren Hsueh, Satoshi Mitarai

## Abstract

*Mycobacterium abscessus* complex (MABC) is an emerging non-tuberculous mycobacterium (NTM). Specific MABC clones are reportedly spreading globally in cystic fibrosis (CF) patients, however, associated genomic epidemiology studies are lacking in East Asia. Analysis of whole-genome sequencing data for MABC isolates from 220 pre-treatment, non-CF patients in Japan and Taiwan revealed that 112/220, 105/220, and 3/220 were *M. abscessus* subsp. *abscessus* (ABS), *M. abscessus* subsp. *massiliense* (MAS), and *M. abscessus* subsp. *bolletii* (BOL), respectively. No significant differences in subspecies composition were noted based on location. Moreover, >50% of ABS and >70% of MAS were related to four predominant clones in the region. Known mutations conferring acquired macrolide resistance were rare (1.4%) and not enriched in the predominant clones. Conversely, the macrolide-susceptible *erm*(41) T28C mutation was significantly enriched in one predominant ABS clone. The most predominant ABS clone was genetically related to the dominant circulating clone (DCC). Hence, we have clarified the relationship between the predominant clones in Japan and Taiwan, and those reported in the international CF patient community. Our results provide insights regarding the genetic characteristics of globally dominant and area-specific strains isolated from patients with or without CF, as well as differences between globally spread and regionally-specific strains.

## Introduction

Non-tuberculous mycobacteria (NTM) is defined as a mycobacterial species other than *Mycobacterium tuberculosis, M. leprae*, and *M. lepromatosis* that causes leprosy. As of August 2021, a total of 194 species have been officially registered in the genus *Mycobacterium* with valid nomenclature (1), many of which cause diseases in mammals, including humans. NTM ubiquitously exists in the environment, including water, soil, plants, and animals. Although the global incidence of tuberculosis has declined gradually, pulmonary infectious diseases caused by NTM are globally emerging and frequently intractable due, in part, to their intrinsic resistance to many antibiotics (2). The *Mycobacterium abscessus* complex (MABC) account for a group of frequently isolated opportunistic pathogens in patients with or without host risk factors, such as cystic fibrosis (CF), bronchiectasis, and other immunocompromised statuses (3–5). The occurrence of pulmonary MABC infection has long been investigated, particularly among CF patients, primarily throughout Europe and the Americas (3, 6, 7), and has also been reported among non-CF patients in some Asian countries, including Japan, South Korea, and Taiwan (8–12).

The MABC is a triad of rapidly growing NTM comprising *M. abscessus* subsp. *abscessus* (ABS), *M. abscessus* subsp. *bolletii* (BOL), and *M. abscessus* subsp. *massiliense* (MAS) (13). The clinical differences among the subspecies regarding the incidence, manifestation, and prognosis are gradually being elucidated and reflect the clinical importance of differences in subspecies (2, 14, 15). With recent advances in whole-genome sequencing (WGS) technologies, a series of epidemiological studies on MABC isolates have been conducted at different scales, including outbreaks within a single hospital (16, 17), nationwide (18, 19), and even those spread intercontinentally (20, 21). Using WGS data of geographically diverse MABC isolated from patients with CF, Bryant *et al*. revealed that most isolates form dense clusters with low genetic diversity, and three dominantly circulating clones (DCCs) were identified, namely, two ABS clones (DCC1 and DCC2) and one MAS clone (DCC3) (20). Subsequent epidemiological studies using WGS at CF centers in other cohorts confirmed the presence of ABS and MAS clones widely distributed among the patients studied (19, 22), however, it is unclear whether these findings apply to MABC isolated from non-CF patients and whether these DCCs are also prevalent in East Asian countries, where few patients are diagnosed with CF (23, 24).

Each subspecies exhibits different susceptibility to macrolides, one of the key antibiotics in the treatment of MABC infections. Of the three subspecies of MABC, nearly all MASs are susceptible to macrolides due to the presence of the truncated unfunctional erythromycin ribosomal methylase (*erm*)(41) gene (25, 26). The remaining two subspecies with the functional *erm*(41) gene induce macrolide resistance via methylation of the target molecule, however, become macrolide-susceptible when the *erm*(41) gene loses its function through T to C substitution at position 28 (27, 28). In addition to intrinsic macrolide resistance, MABC can acquire mutational macrolide resistance through substitutions on the *rrl* gene encoding the 23S rRNA (29, 30). Since these mutations likely impact long-term treatment and promote poor outcomes of MABC infection compared to other NTM diseases (31), detailed epidemiological analyses of macrolide resistance-associated mutations are required to control MABC infection. However, the incidence of these mutations in MABC populations derived from non-CF untreated patients in Asian countries, as well as the relationship between these mutations and the predominant clones, are poorly understood.

In this study, we sequenced the genomes of 220 MABC clinical isolates, all of which were obtained from non-CF patients before treatment in four hospitals in three East Asian locations (Tokyo, Okinawa, and Taiwan) covering subtropical to temperate climate zones over five years. Using the data set, we examined subspecies distribution, the incidence of macrolide resistance-associated mutations, as well as prevalent clones and their genetic features to determine the epidemiological distribution of MABC in Japan and Taiwan.

## Results

### Isolation of MABC subspecies in Japan and Taiwan

We first identified subspecies of 220 MABC clinical isolates based on WGS data to examine the subspecies distribution in Japan and Taiwan. In a maximum-likelihood phylogenetic tree, isolates were divided into three major clades corresponding to each of the three subspecies of MABC (Fig. 1). This result was confirmed by calculating the average nucleotide identity (ANI) values among the MABC isolates. The minimum ANI among MABC clinical isolates was 96.4%, all ANI values within the three subspecies were > 98% (minimum: 98.1%), while all ANI values between subspecies were < 98% (maximum: 97.6%; Fig. S1). This indicates that the species and subspecies boundaries of the MABC clinical isolates were approximately 96% and 98% ANI, respectively, which is consistent with previous results (15). Of the 220 isolates, 112 (50.9%), 105 (47.7%), and 3 (1.4%) were identified as ABS, MAS, and BOL, respectively (Fig. 1). We detected no significant difference between the composition of MABC subspecies in Tokyo, Okinawa, and Taiwan (Fig. 1, Wald test).

**Figure 1.**
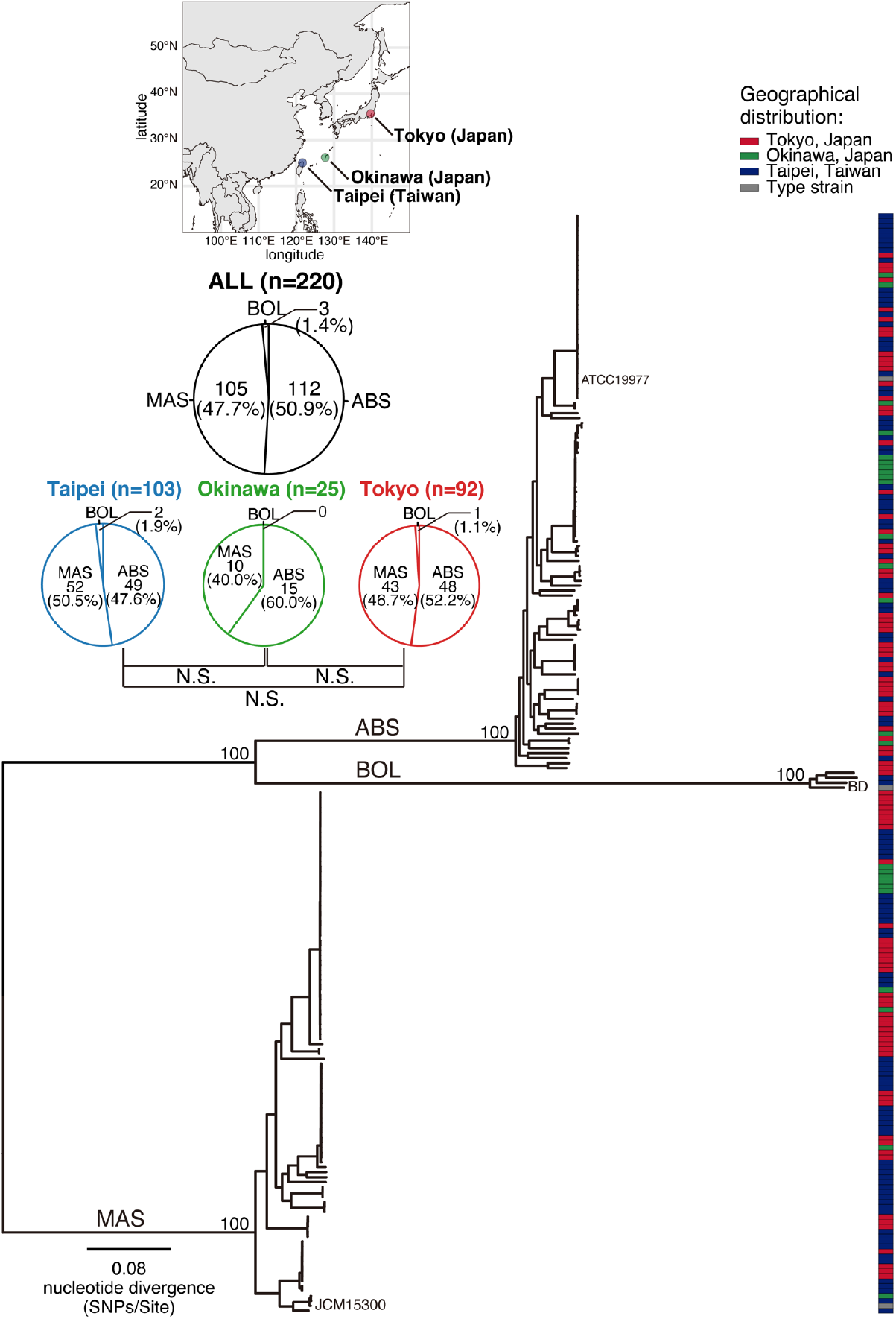
Phylogeny of 220 clinical isolates of MABC in Japan and Taiwan. Core-genome alignment of 220 isolates and three reference strains (ABS ATCC19977, MAS JCM15300, and BOL BD) of MABC was generated. A complete genome sequence of ATCC19977 was used as a reference. An alignment containing 235,540 recombination-free variable positions was used with RAxML to estimate a maximum-likelihood tree with 300 bootstrap replicates. Bootstrap values for the major nodes are shown. Scale bar indicates the mean number of nucleotide substitutions per site (SNPs/Site) on the respective branch. The red, green, and blue boxes indicate locations where each clinical isolate was obtained, and the gray box indicates the reference strain (ATCC19977). Pie charts indicate the ratio of the three subspecies of isolates identified in all (n=220), Tokyo (Japan, n=92), Okinawa (Japan, n=25), and Taipei (Taiwan, n=103), respectively.

### Prevalent clones of ABS and MAS in Japan and Taiwan

To investigate the genetic relatedness of isolates from individual patients in Japan and Taiwan, we analyzed the phylogeny of ABS and MAS separately (Fig. 2, Fig. 3). Among the 112 ABS isolates, we identified six clusters (ABS-EA1 to ABS-EA6, shown in Fig. 2), including 85 isolates (75.9%). Of these isolates, 37 (33.0%) and 25 (22.3%) from the three locations belonged to the largest clusters (ABS-EA1 and ABS-EA2, respectively) (Fig. 2). The other three ABS-EA clusters (ABS-EA3, 5, and 6) consisted of isolates from two different sites (Tokyo/Okinawa, Tokyo/Taipei, or Taipei/Okinawa). We identified five clusters of 105 MAS isolates, including 87 isolates (82.9%) (MAS-EA1 to MAS-EA5, shown in Fig. 3). Of these clustered isolates, 51 (48.6%) and 20 (19.0%) from the three locations belonged to the first (MAS-EA1) and second (MAS-EA2) largest clusters (Fig. 3), respectively, while the other three clusters (MAS-EA3, 4, and 5) consisted of isolates from two locations. We also examined the population differences between ABS and MAS in this region. The genome-wide nucleotide diversity of MAS was significantly higher than that of ABS (Fig. S2A, *p* < 5.1e-06, two-sided Wilcoxon rank-sum test), and the total number of genes within MAS isolates was larger than that of the ABS isolates (Fig. S2B), whereas the pairwise genetic distances between isolates within each of the MAS-EA clusters were significantly lower than those of the ABS-EA clusters (Fig. S2C, *p* < 2.2e-16, two-sided Wilcoxon rank-sum test).

**Figure 2.**
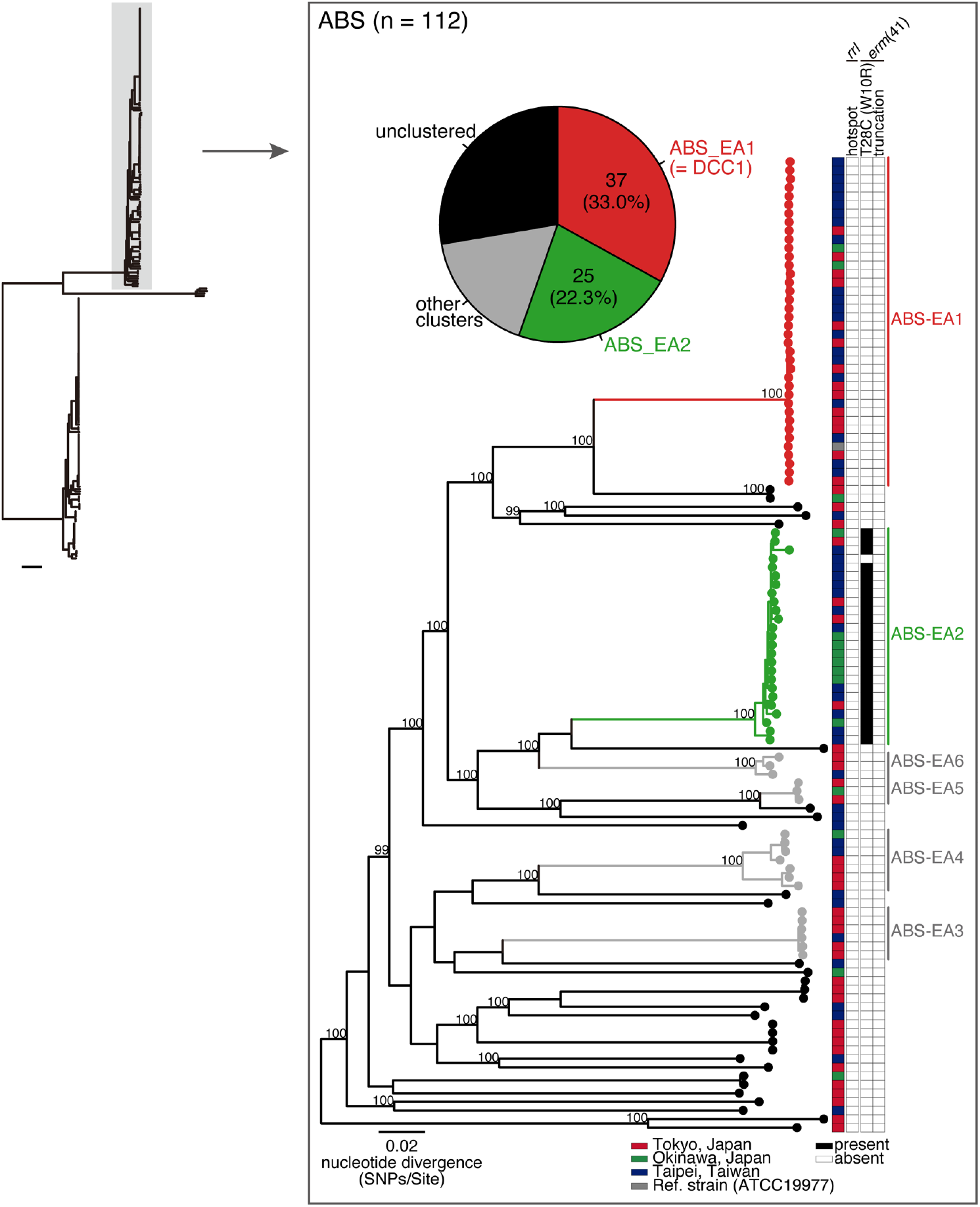
Clustering analysis of ABS in Japan and Taiwan, and mutations associated with inducible or acquired macrolide resistance. A core-genome alignment of 112 ABS clinical isolates and a reference strain ATCC 19977 was generated (=3,963,788 bp, covering 78.2% of the reference genome). The alignment containing 76,114 recombination-free variable positions within the core-genome was used to estimate a maximum-likelihood tree with 300 bootstrap replicates. Bootstrap values > 98% for the major nodes are shown. Six monophyletic clusters (ABS-EA1 to ABS-EA6), identified by TreeGubbins, are shown. The pie chart indicates the ratio of identified clusters, and two dominant clusters (ABS-EA1 and ABS-EA2) are depicted in red and green. The location where each clinical isolate was obtained (as shown in Fig. 1). The presence (black) and absence (white) of macrolide resistance-associated mutations are indicated. Scale bar; the mean number of nucleotide substitutions per site (SNPs/Site) on the respective branch.

**Figure 3.**
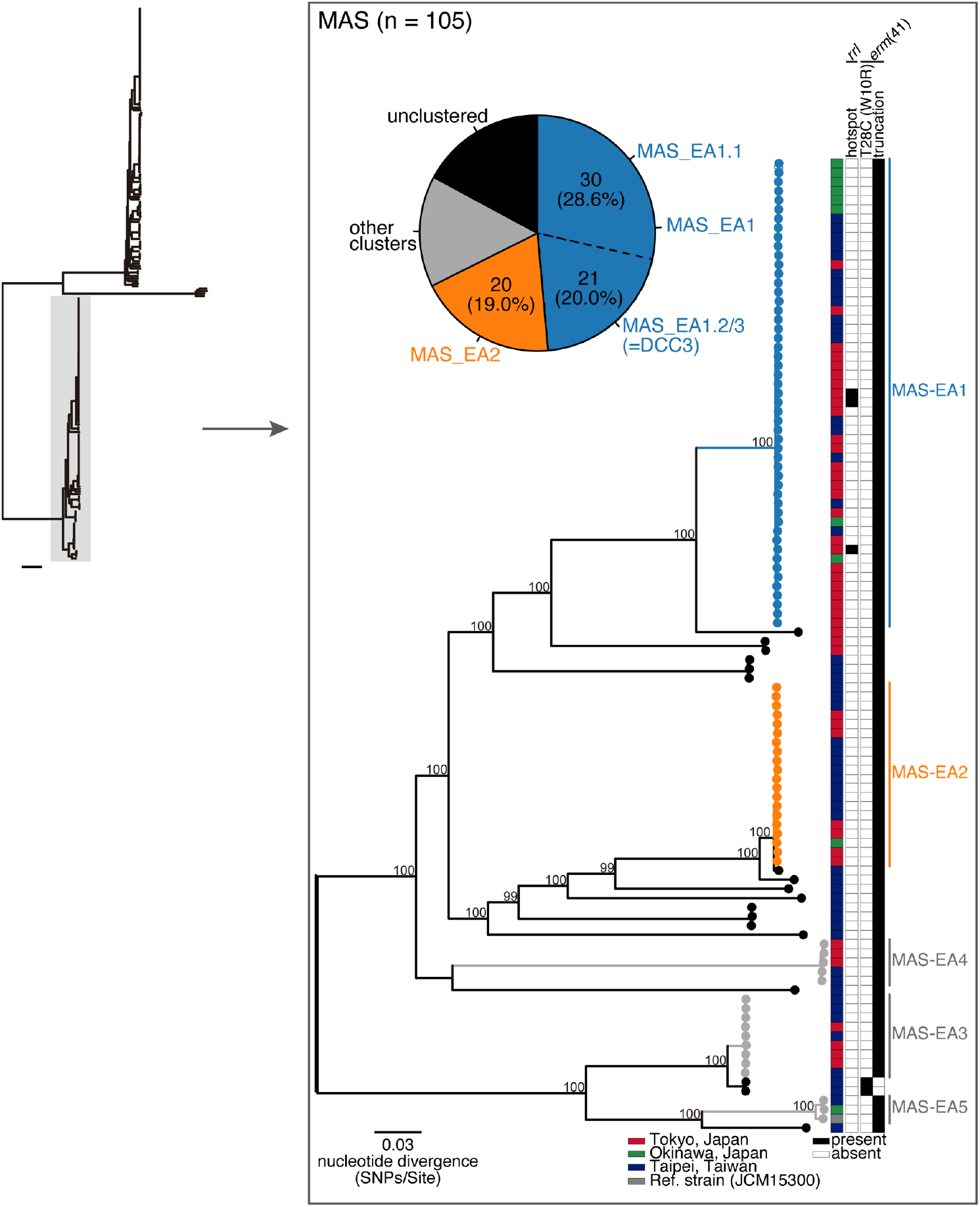
Clustering analysis of MAS in Japan and Taiwan, and mutations associated with inducible or acquired macrolide resistance. A core-genome alignment of 105 MAS clinical isolates and a reference strain JCM15300 was generated (=4,033,769 bp, covering 81.0% of the reference genome). The alignment containing 48,718 recombination-free variable positions located within the core-genome was used to estimate a maximum-likelihood tree with 300 bootstrap replicates. Bootstrap values > 98% for the major nodes are shown. Five monophyletic clusters (MAS-EA1 to MAS-EA5), identified by TreeGubbins, are shown. The pie chart indicates the ratio of identified clusters and two dominant clusters (MAS-EA1 and MAS-EA2) depicted in blue and orange. The location of each clinical isolate, and the presence (black) and absence (white) of macrolide resistance-associated mutations were indicated in Fig. 2. Scale bar; the mean number of nucleotide substitutions per site (SNPs/Site) on the respective branch.

### Mutations associated with macrolide resistance of ABS/MAS in Japan and Taiwan (East Asia, EA)

Since macrolides, such as clarithromycin and azithromycin, are first-line antibiotics for the treatment of MABC infection, it is important to determine the incidence of genetic mutations associated with macrolide susceptibility in the presence of circulating clones in Japan and Taiwan. We found that no ABS isolates harbored mutations in the *rrl* hotspot (2269 to 2271 bp in the *rrl* gene of ATCC19977) (Fig. 2). Among the 112 ABS, 24 (21.4%) carried the *erm*(41) T28C sequence variant. Notably, all *erm*(41) T28C sequences belonged to the ABS-EA2 and exhibited significant enrichment in this clade (Fig. 2, *p* < 2.2e-16, Fisher’s exact test). Three of the 105 MAS (2.9%) had substitutions in the hotspot on the *rrl* gene, all of which belonged to MAS-EA1, however, were not significantly enriched (Fig. 3, Fisher’s exact test). Nearly all MAS (103 isolates) had truncation of the *erm*(41) gene without substitution in the hotspot of the *rrl* gene. However, two MAS (TWN-024 and TWN-041) displayed a full-length *erm*(41) gene, with a simultaneous T28C substitution. A total of 102 MAS isolates (97.1%) were predicted to be susceptible to macrolides. Together with the fact that no BOL (n = 3) displayed substitutions in the *rrl* hotspot or T28C mutation in the *erm*(41) gene (data not shown), 126/220 MABC isolates (57.3%, 24 ABS and 102 MAS) were predicted to be susceptible to macrolides due to dysfunctional *erm*(41), whereas 91 isolates (41.4%, 88 ABS and 3 BOL) may induce macrolide resistance, and the remaining three isolates (1.4%) carried mutations that have been reported to confer macrolide resistance.

### The relationship between ABS prevalent in Japan and Taiwan and previously described prevalent clones worldwide

We next investigated the relationships between prevalent clones in Japan and Taiwan and previously described DCCs. Publicly available raw sequence data for 349 ABS from individual CF patients in seven countries (one isolate per patient, all additional isolates are listed in Table S3) (20) were combined with our data set. In the ABS population, we identified 16 clusters (ABS-GL1 to ABS-GL16 shown in Fig. 4), of which 11 (ABS-GL1, 2, 3, 4, 5, 7, 8, 11, 14, 15, and 16) consisted of isolates from more than two regions (Fig. 4). Two clusters (ABS-GL10 and ABS-GL13) exclusively consisted of isolates from Japan and Taiwan, suggesting the existence of specific clones in the East Asian region. All ABS isolates belonging to ABS-EA1 were clustered with isolates that were identified as DCC1 (=Absc1) within the global CF patient community (20) (Fig. 4). Although all ABS belonging to DCC2 (=Absc2) (20) were clustered in ABS-GL2, no isolate from Japan and Taiwan belonged to this cluster (Fig. 4). We also found that all isolates belonging to ABS-EA2 clustered in ABS-GL3 with several Absc clusters. Notably, 97 out of 114 isolates (85.1%) belonged to the clade carrying T28C in the *erm*(41) gene, whereas the remaining 17 isolates did not (Fig. 4). Although isolates belonging to this clade exclusively displayed the substitution, it appeared that isolates of several sub-clades within ABS-GL2 reversed to the wild-type T28 genotype in the *erm*(41) gene (Fig. 5A, Fig. S3). To explore the mode of reacquisition of the T28 genotype, we examined the mutation pattern of the *erm*(41) gene in all ABS clinical isolates (Fig. 5, Fig. S3). We found that the ancestral form of *erm*(41) in clinical isolates belonging to the ABS-GL2 carries the T159C, A238G, and A330C sequence variants in addition to T28C (Fig. 5A, Fig. S3). However, isolates belonging to the four sub-clades and one isolate (TWN-080) that reversed the T28 genotype, showed distinctly different mutation patterns (red circles in Fig. 5A). In these isolates, we detected that approximately 1.4 to 52 kbp fragments harboring the wild-type *erm*(41) T28 genotype were inserted by recombination around the *erm*(41) gene (red and blue shaded boxes in Fig. 5A, Fig. 5B). We also observed the phylogenetically high recombination potential of the ABS-GL2 isolates compared to that of other clinical isolates (Fig. S3). In contrast, *erm*(41) from an isolate (RVI3T, blue circles in Fig. 5A) displayed the same mutation pattern as the ancestral type, with the exception of wild-type T28, and no recombination event was detected in this genomic region of RVI3T (Fig. 5A, B).

**Figure 4.**
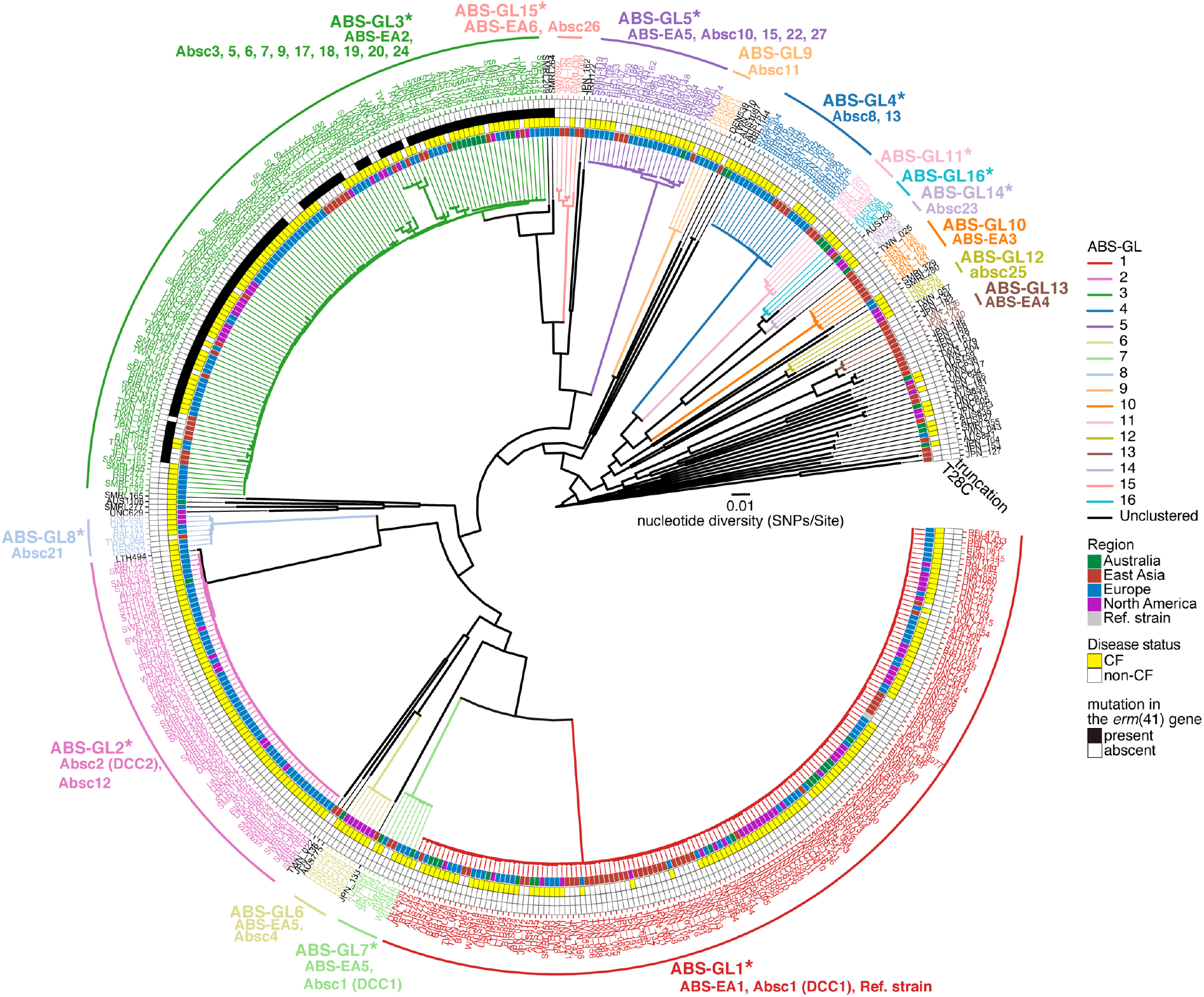
The phylogenetical relationship between ABS in Japan and Taiwan, and that in other countries. Phylogeny of ABS from several countries was estimated using a core-genome alignment of 461 clinical isolates from four regions (Australia, East Asia, Europe, or North America). The complete genome sequence of ATCC19977 was used as a reference. The alignment containing 102,613 recombination-free variable positions located in the core-genome (3,584,621 bp, covering 70.7% of the reference genome) was used with RAxML to construct a maximum-likelihood tree with 300 bootstrap replicates. The 16 monophyletic clusters (ABS-GL1 to ABS-GL16), identified by TreeGubbins, and corresponding clusters identified in Fig. 2 and by Bryant *et al*. (2016) are shown. The presence (black) and absence (white) of inducible macrolide resistance-associated mutations in the *erm*(41) gene are indicated. Disease status (CF; yellow and non-CF; white) of corresponding patients are shown. Each color box corresponds to the region where the clinical isolate was obtained. Asterisks indicate clusters consisting of isolates from more than two regions. Scale bar indicates the mean number of nucleotide substitutions per site (SNPs/Site) on the respective branch.

**Figure 5.**
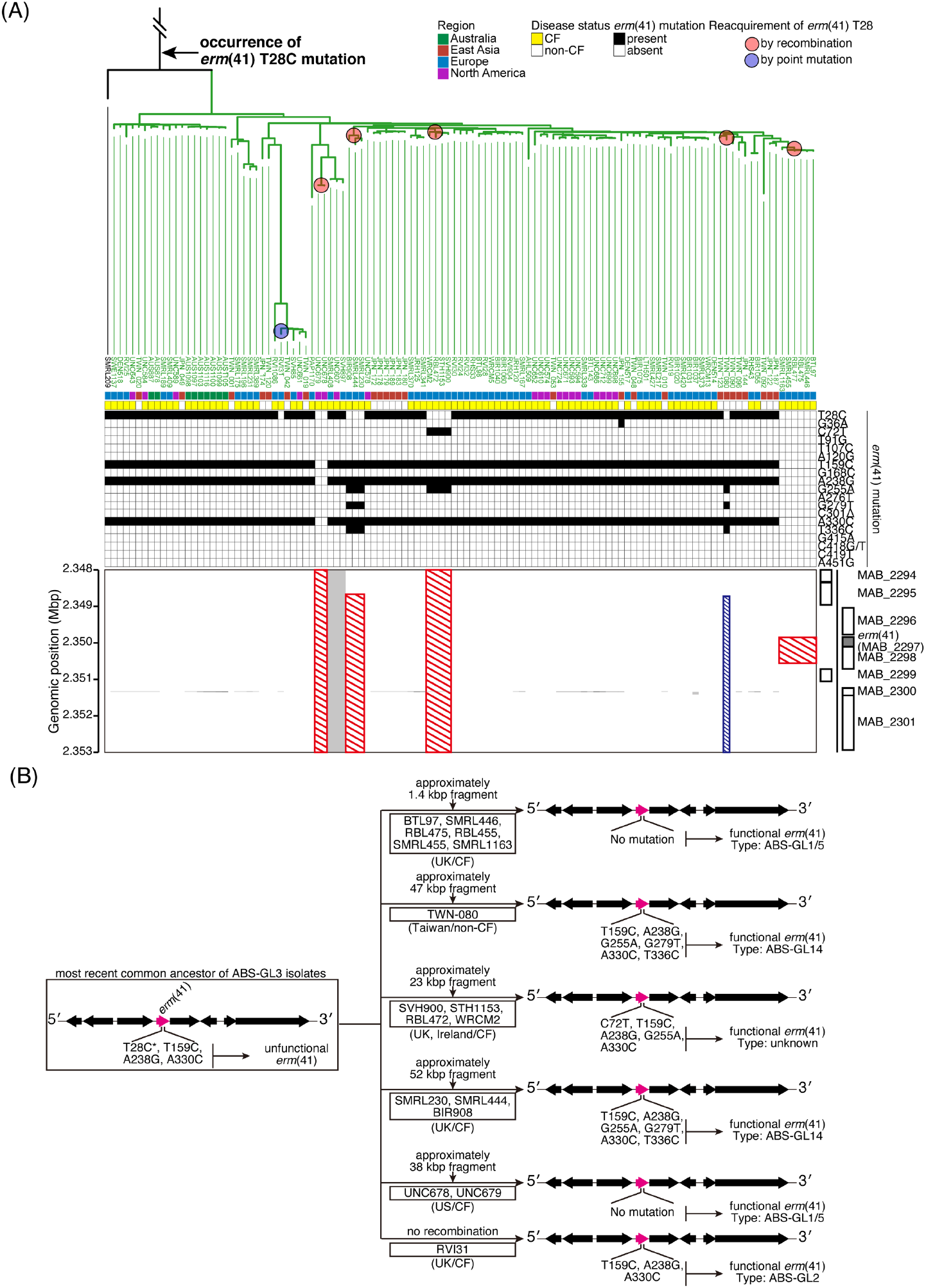
Reacquisition of wild-type T28 genotype in *erm*(41) of ABS clinical isolates belonged to the ABS-GL2 cluster. (A) Recombination events in the genomic region around the *erm*(41) gene of ABS clinical isolates belonged to the ABS-GL2 cluster. Red or blue shaded boxes indicate common or sporadic recombination events. The presence and absence of mutations in the *erm*(41) gene, disease status (CF or non-CF) of corresponding patients, and the origin of the clinical isolate are shown in Fig. 4. (B) A schematic depiction regarding the reversion of the wild-type *erm*(41) T28 genotype. Black and magenta arrows indicate genes, the fourth of which is *erm*(41).

### The relationship between MAS prevalent in Japan and Taiwan (EA) and previously described prevalent clones worldwide

In addition to ABS isolates, we assembled publicly available WGS data for 127 MAS clinical isolates from CF patients from seven countries (listed in Table S3) (16, 20, 32) and combined them with our data set. We identified 11 clusters (MAS-GL1 to MAS-GL11 shown in Fig. 6), of which eight (MAS-GL1, 2, 3, 4, 6, 8, 9, and 11) could be considered as globally dispersed clones, while the remaining three were specific to the target region (MAS-GL5, 7, and 10 shown in Fig. 6). MAS-GL2 contained all isolates belonging to MAS-EA2, which is the second-largest cluster in Japan and Taiwan (Fig. 3), and is described as an internationally spreading clone Mass5 (20). Although TWN-024 and TWN-041, carrying full-length *erm*(41), were clustered with DEN526 (from Denmark) and UNC618 (from the USA) in the MAS-GL8, the latter two isolates showed truncation of the *erm*(41) gene, as well as all other MAS isolates (Fig. 6). The mutation pattern of the *erm*(41) gene in TWN-024 and TWN-041 was similar to that in ABS isolates belonging to ABS-GL2 (T28C, T159C, A238G, and A330C sequence variants, data not shown). We also found one isolate (JRH124 from the UK) that did not belong to any MAS-GL clusters, and sporadically displayed full-length *erm*(41) without T28C substitution (Fig. 6). MAS-GL1 contained all isolates belonging to MAS-EA1, DCC3 (Mass1), and Mass3 (20) (Fig. 6, Fig. 7A). It should be noted that MAS-GL1 contained isolates from patients during outbreaks in two CF centers (16, 32) (magenta boxes in Fig. 6).

**Figure 6.**
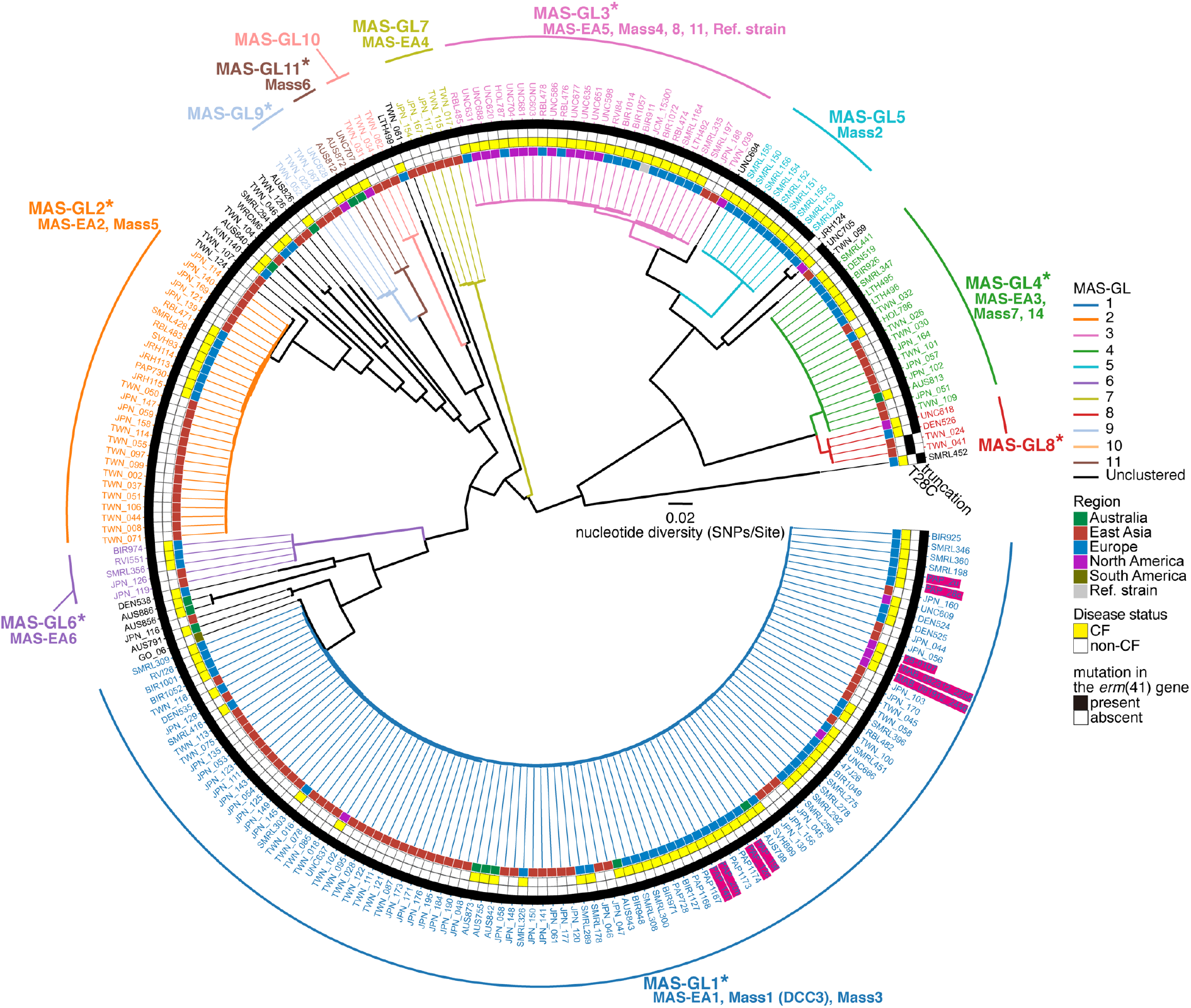
The phylogenetic relationship between MAS in Japan and Taiwan, and that in other countries. Phylogeny of global MAS was estimated using a core-genome alignment of 233 clinical isolates from five regions (Australia, East Asia, Europe, North America, and South America). The complete genome sequence of JCM15300 was used as a reference. The alignment containing 58,975 recombination-free variable positions located in the core-genome (3,789,583, covering 76.1% of the reference genome) was used with RAxML to construct a maximum-likelihood tree with 300 bootstrap replicates. The 11 monophyletic clusters (MAS-GL1 to MAS-GL11) identified by TreeGubbins with corresponding clusters identified in Fig. 3 and by Bryant *et al*. (2016) are shown. The presence and absence of mutations in the *erm*(41) gene, disease status (CF or non-CF) of corresponding patients, and origin of the clinical isolate are shown in Fig. 4. Asterisks indicate clusters consisting of isolates from more than two regions. Magenta boxes indicate clinical isolates that caused nosocomial outbreaks of MAS (16, 30). The scale bar indicates the mean number of nucleotide substitutions per site (SNPs/Site) on the respective branch.

**Figure 7.**
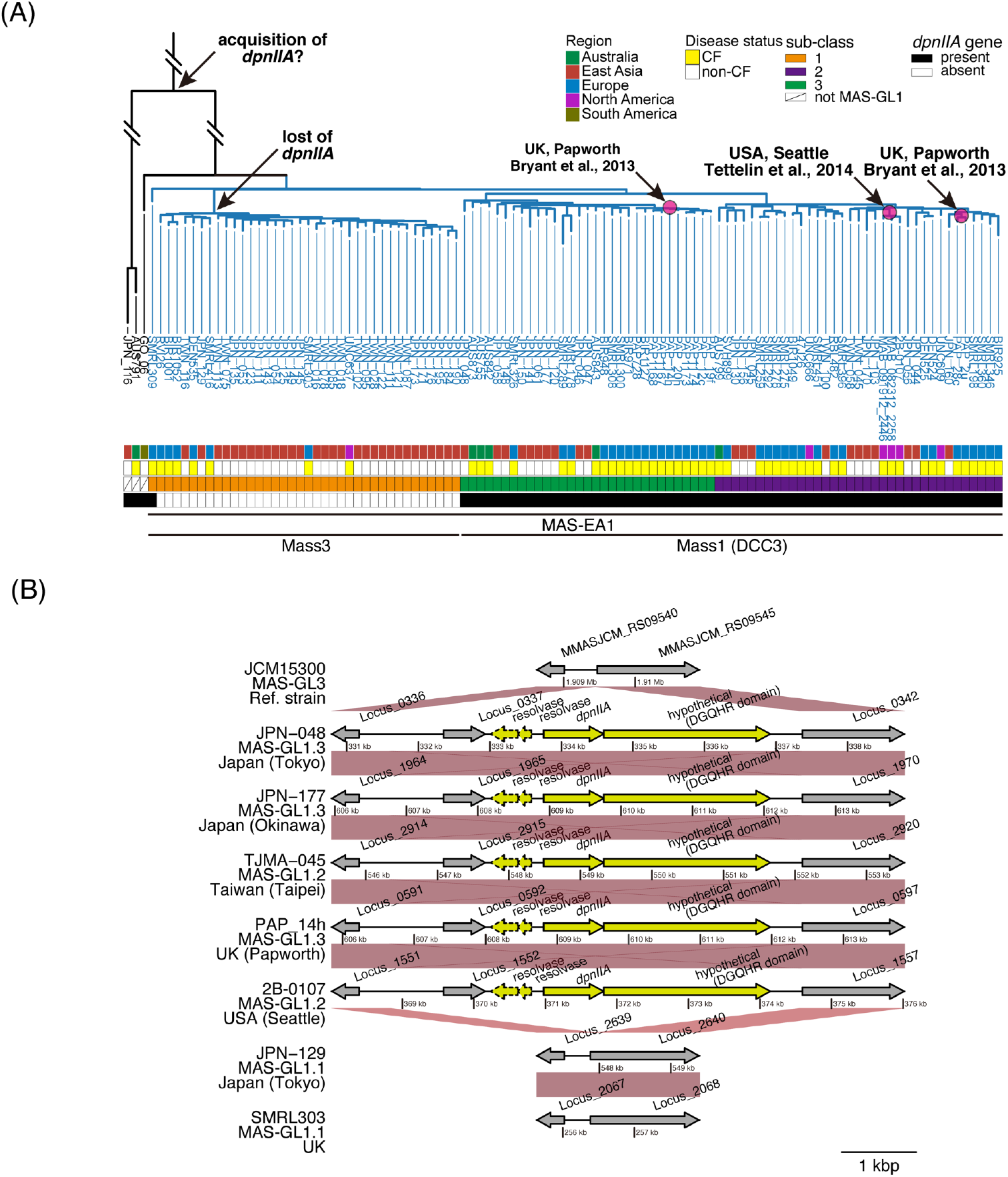
Genetic components exclusively associated with MAS clinical isolates that were closely related to previously reported outbreak strains. (A) Sub-cluster analysis of MAS belonged to the MAS-GL1 cluster. The phylogenetic tree of MAS clinical isolates was estimated as described in Fig. 6. Sub-clusters were inferred using fastBAPS (56) with default settings. Herein, the core-genome alignment of clinical isolates, which belonged to the MAS-GL1 cluster, was used as an input file. The presence (black) and absence (white) of the *dpnIIA* gene, disease status (CF or non-CF) of corresponding patients, and the region where the clinical isolate was obtained are shown. (B) Acquisition examples of the *dpnIIA* locus between clinical isolates belonged to the MAS-GL1.2 and MAS-GL1.3 clusters. Arrows indicate genes annotated with DFAST-core (53) and dotted arrows indicate pseudogenes estimated using DFAST-core. Orthologous genes between clinical isolates are shown with red connections and were plotted using genoPlotR (57). Five genes exclusively associated with clinical isolates that belonged to the MAS-GL1.2 and MAS-GL1.3 clusters are indicated in yellow.

In our sample set, half of the MAS isolates belonged to MAS-GL1 (MAS-EA1). These observations prompted us to further investigate MAS-GL1 phylogeny. We identified three sub-clusters within the MAS-GL1 (MAS-GL1.1 to MAS-GL1.3, Fig. 7A) and found that DCC3 (Mass1) isolates, as well as isolates from patients during previous outbreaks, exclusively belonged to MAS_GL1.2 or MAS-GL1.3 (Fig. 7A). In MAS clinical isolates from Japan and Taiwan, 21 out of 105 (20%) belonged to MAS_GL1.2 or MAS-GL1.3, while 30 (28.6%) belonged to MAS-GL1.1 (Fig. 3). To genetically characterize MAS isolates that belonged to MAS_GL1.2 or MAS-GL1.3, we identified four genes that were significantly associated with these clades (*p* < 2.7e-52 after Bonferroni correction, Fisher’s exact test, Table S4). These genes included two resolvases, both of which may be unfunctional due to frameshift, a DGQHR domain-containing hypothetical protein, and a site-specific DNA-adenine methyltransferase *DpnIIA* (Fig. 7B). We also found that the most recent common ancestor (MRCA) of MAS-GL1 retained the *DpnIIA* gene, and a descendant lineage leading to DCC3 also retained this gene, meanwhile, another descendant lineage leading to Mass3 was determined to have lost this gene.

## Discussion

Herein, we investigated the epidemiological distribution of pulmonary MABC infection in Japan and Taiwan based on WGS data from a set of clinical isolates obtained from non-CF pulmonary MABC patients living in three locations (Tokyo, Okinawa, and Taipei). As such, this serves as the first WGS-based epidemiological study focusing on MABC clinical isolates from non-CF patients in East Asia, thus, expanding the previous findings reported for CF patients in Western countries (16–22, 33). Our sample set, all of which were obtained from patients diagnosed with pulmonary MABC disease, consisted of approximately half ABS, while the other half were MAS, and a small number of BOL. No geographical differences were noted in the subspecies composition of the three locations examined, which agrees with the results of previous epidemiological studies on non-CF patients conducted by conventional methods in Japan and other East Asian cohorts (12, 34, 35). While the incidence of BOL was very low in our cohort, BOL reportedly accounts for 10–20% of MABC in European cohorts of CF patients (22, 36). Although the composition of the three subspecies is difficult to compare directly, as it primarily depends on the clinical and environmental settings of each institution, these observations may suggest that there are differences in the geographic distribution of BOL and/or the susceptibility to BOL between CF and non-CF patients.

Since macrolide antibiotics are key drugs for the treatment of pulmonary MABC infection, we revealed the incidence of macrolide resistance-associated mutations in our sample set. Although all of our clinical isolates were derived from untreated patients, 1.4% of the isolates, all of which were identified as MAS, were expected to show macrolide resistance due to mutations in the *rrl* gene, and these mutations did not enrich any dominant clones of MABC in Japan and Taiwan. Our genotyping analysis suggested that at least 57% of MABC clinical isolates were susceptible to macrolides due to the loss of function of the *erm*(41) gene, suggesting that macrolide-containing regimens are effective in treating more than half of MABC infections in our cohort. Notably, over 20% of ABS carried the macrolide-susceptible *erm*(41) T28C mutation, and this mutation was significantly enriched in one of four dominant clones of MABC in this area. Although it is currently unknown whether the ABS *erm*(41) T28C sequevar is present in similar proportions elsewhere, these results emphasize the clinical importance of accurate genotyping of the *rrl* and *erm*(41) gene before initiation of MABC treatment, as recommended by recent treatment guidelines for pulmonary MABC infection (14, 15).

There were several clusters of isolates from both Japan and Taiwan, but the number of isolates in each cluster differed. In ABS isolates, 33% and 22% of isolates belonged to the largest and second-largest clusters (ABS-EA1 and ABS-EA2), respectively, representing dominant clones in Japan and Taiwan. Remarkably, the ABS *erm*(41) T28C mutation, which is susceptible to macrolide antibiotics (37), exclusively accumulated in the ABS-EA2 cluster. These results indicate that the ABS *erm*(41) T28C sequevar can be considered as one of the major clades in Japan and Taiwan. As for MAS, this trend was more pronounced, with more than 70% of MAS isolates belonging to two large clusters (MAS-EA1 and MAS-EA2). These results suggest that specific clones of ABS and MAS are intensively isolated from non-CF patients in Japan and Taiwan. A comparison of the population structure of the two MABC subspecies revealed that MAS has higher nucleotide diversity and a larger gene repertoire, but shows lower genetic distances among isolates belonging to the identified clusters. These results suggest that MAS is a more genetically diverse population than ABS, and certain homologous clones may adapt to preferentially infect the respiratory system.

Bryant *et al*. reported that two ABS clones (DCC1 and DCC2) and one MAS clone (DCC3) are dominantly spreading worldwide (20). Hence, we analyzed whether these dominant clones also prevail in non-CF patients in East Asian regions. Our results indicate that clinical isolates related to DCC1 clones are also common in Japan and Taiwan. However, no clinical isolates related to DCC2 clones were identified in our sample set, and clinical isolates related to DCC3 clones were not necessarily dominant in our sample set. Consistent with our results, recent genomic epidemiological studies have also reported that DCC1 clones are predominant in CF cohorts across the US, whereas DCC2 and DCC3 are not (19, 22). Thus, it is suggested that the DCC1 clone is distributed worldwide and preferentially infects, and causes pulmonary MABC disease, over other clones in both CF and non-CF patients; however, it remains unclear whether DCC2 or DCC3 preferentially causes disease, compared to other clones. We also identified several regionally-specific, and globally spreading clones. These observations imply that some MABC clones can spread internationally, while others do not. Further genomic, epigenomic, and phenotypic analyses are needed to understand the differences in pathogenicity, as observed in *Mycobacterium tuberculosis* (38–40).

It should be noted that the ABS *erm*(41) T28C isolates from several countries belonged exclusively to ABS-GL2, indicating that this sequence variant can be considered as one of the globally spreading clones. However, in a previous study (20), the *erm*(41) T28C sequevar was divided into several minor clusters and was not recognized as a single cluster (Fig. 4). This is perhaps because our phylogenetic trees were estimated using the core-genome region without recombination sites, whereas Bryant *et al*. included recombination sites in their estimation of the phylogenetic tree (20). Since the ABS *erm*(41) T28C isolates caused more recombination events than other ABS isolates (Fig. S3), we presume that recombination had a relatively large effect on the phylogenetic tree estimation and clustering of the ABS *erm*(41) T28C isolates (41). Notably, some isolates within ABS-GL2 appeared to reverse the wild-type T28 genotype in the *erm*(41) gene. We identified two patterns of reversion of the wild-type T28 genotype. The first is the reversion of the wild-type *erm*(41) T28 genotype by point mutation, while the second involved the incorporation of the *erm*(41) gene from isolates belonging to other clusters into isolates belonging to ABS-GL2. It was also observed that isolates belonging to ABS-GL2 showed high recombination potential compared to those belonging to other clades. Since there are no reports that the *erm*(41) T28C mutation is detrimental to the survival of the bacterium in macrolide-free environments, one possible explanation for the “genetic throwback” is that the isolates belonging to ABS-GL2 have undergone adaptive evolution to survive in the macrolide environment within the host and that the isolates may be infecting other patients indirectly or directly (42). We also observed two MAS isolates carrying the full-length *erm*(41) genes with the T28C mutant and one MAS isolate carrying the full-length *erm*(41) gene. These results suggest that horizontal transfer of the *erm*(41) gene that alters macrolide susceptibility may occur, although not frequently, between ABS and MAS.

In the region of this study, more than half of the MASs belonged to MAS-GL1. This large cluster was divided into three sub-clades, two of which (MAS-GL1.2 and MAS-GL1.3) were exclusively related to isolates from patients during outbreaks in two CF centers in the USA and UK (16, 32), and were annotated as DCC3 (20). Thus, we searched for genes that were significantly associated with MAS-GL1.2 and MAS-GL1.3. The genes included a functional methyltransferase *dpnIIA* gene, which is expected to belong to the “DpnII restriction gene cassette.” DpnII only methylates double-stranded DNA, whereas DpnI can methylate both single- and double-stranded DNA, although both enzymes act at 5’-GATC-3’ in DNA (43). A recent study demonstrated that the *dpnIIA* gene of MABC participates in altering the genome-wide methylation and expression levels of several genes, and is required for intracellular survival (42). Although other mycobacterial species, such as *Mycobacterium* spp. QIA-37 and *Mycobacterium* sp. TRM80801, express the homologue (69.4% and 55.8% identity in amino acid sequence, data not shown), there is no other available functional information on the gene. Note that the ratio of isolates related to DCC3 in our cohort was not necessarily higher than those related to other MAS clades. This result is consistent with previous genomic epidemiological studies using a set of clinical isolates from patients with CF (19, 22). In addition, strains belonging to MAS-GL1.1 and MAS-GL2 (=MAS-EA2, Mass5), which lost the *dpnIIA* gene, were isolated to the same extent as isolates related to DCC3. These results indicate that, at least in our cohort, the *dpnIIA* gene alone does not serve as a determinant of whether a MABC clone is dominant. Nevertheless, all strains isolated during nosocomial outbreaks in CF centers (16, 32) carry the *dpnIIA* gene. It is also noteworthy that a strain (GO-06) isolated from a nosocomial infection at the surgical site of several patients in Brazil (44) still carries the *dpnIIA* gene, although it is not related to DCC3 (Fig. 7). Further research is needed to determine how the acquisition of the *dpnIIA* gene led to the nosocomial outbreak of MAS, and whether there are other cases of outbreaks caused by clade-specific gene acquisition, to understand the pathogenic evolution of MABC.

The present study has several limitations. First, this study was conducted using MABC clinical isolates from only four facilities in three locations; therefore, further validation using other sample sets is needed to examine whether it captures the complete picture of pulmonary MABC disease in Japan and Taiwan. Second, there is a lack of information regarding epidemiological links among patients. Analysis of the genetic distances between clinical isolates from individuals showed the possibility of direct or indirect transmission of MABC (Fig. S2C) (16, 17). Further analysis, including tracking of epidemiological linkages among patients and environmental sampling, is required to understand the mode of transmission of MABC. Finally, as there are few CF patients in Asia, our sample set did not include clinical isolates derived from CF patients in Japan and Taiwan. Therefore, it cannot be determined whether the detected area-specific clusters were caused simply by differences in the geographical distribution of MABC or by differences in patients’ disease status (CF or non-CF).

In conclusion, we revealed prevalent clones and the incidence of macrolide resistance-associated mutations in MABC isolated from non-CF patients in Japan and Taiwan. We also clarified the relationship between these clones and previously described dominant clones in the international CF patient community. Our results provide a cornerstone for WGS-based epidemiological analysis of pulmonary MABC disease in the East Asian region. Furthermore, it would be helpful for future studies to elucidate the genetic differences between globally predominant and area-specific clones isolated from patients with and without CF.

## Methods

### Sample collection and DNA sequencing

A total of 220 clinical isolates (one isolate per patient before the start of treatment) were recovered from respiratory specimens from 2012 to 2017 (Table S1). We confirmed that all patients met the diagnostic criteria for ATS/ERS/ESCMID/IDSA (14). Of these clinical isolates, 117 were obtained from three hospitals in Japan (Fukujuji Hospital, Keio University Hospital, and Okinawa Chubu Hospital), and 103 isolates were obtained from a hospital in Taiwan (National Taiwan University Hospital). All isolates were classified as MABC by DDH Mycobacteria (Kyokuto Pharmaceutical Industrial, Tokyo, Japan) and/or MALDI-TOF MS (Bruker Daltonics, MA, USA), and were sub-cultured on 2% Ogawa egg slants or 7H10 agar plates supplemented with 10% OADC (BD BBL, MD, USA).

The genomic DNA of each isolate was extracted and purified using a standard enzyme/organic solvent method (45). A WGS library was constructed using the QIAseq FX DNA Library Kit (QIAGEN, Hilden, Germany). Sequencing was performed on the standard NextSeq 550 platform (Illumina Inc., CA, USA) using the NextSeq Mid Output Kit 2.5 (300 cycles, Illumina Inc.) with 150-mer paired-end short reads according to the manufacturer’s instructions. All raw read data of newly sequenced strains in this study (220 isolates) were deposited into the DNA Data Bank of Japan (DDBJ) and the National Centre for Biotechnology Information (NCBI) under BioProject accession number PRJDB10566.

### Phylogenetic analyses based on core-genome variable positions of MABC isolates

The raw read data of each isolate were *de novo* assembled in the Shovill pipeline (https://github.com/tseemann/shovill) with default settings, and contigs were produced without scaffolding. The number of contigs, N50 values, and raw coverage of each isolate are listed in Table S2. The data were combined with the publicly available complete genome sequence of *M. abscessus* subsp. *abscessus* ATCC19977 (46), *M. abscessus* subsp. *massiliense* JCM15300 (47), and *M. abscessus* subsp. *bolletii* BD (48). For phylogenetic analyses, we first performed pairwise genome alignment between ATCC19977 and one of the clinical isolates using the MUMmer package (49) to identify conserved genomic regions among these isolates and the SNP sites located within these regions. We then combined all alignments into a whole-genome alignment, in which each position corresponded to that of the complete genome of ATCC19977 using custom Perl scripts. We identified 3,638,122 bp of the ATCC19977 genome that was conserved among all clinical isolates (core-genome) and identified 235,540 variable positions within the core-genome. This alignment was used to infer recombination sites using Gubbins (50), and 8,358 recombinogenic sites were detected and removed from the alignment. The resulting alignment containing 240,429 recombination-free variable positions located in the core-genome region were used to reconstruct a maximum-likelihood tree using RAxML ver. 8.2.12 (51) with the General Time Reversible (GTR)-GAMMA model of nucleotide substitution and 300 bootstrap replicates. To confirm phylogeny-based subspecies identification, ANI values among all MABC isolates were calculated using fastANI with default settings (52). Multiple whole-genome alignments were used to investigate the genotypes of the *erm*(41) gene (MAB_2297, 2,345,955 bp to 2,346,476 bp in ATCC19977) and the *rrl* gene (MAB_r5052, 1,464,208 bp to 1,467,319 bp in ATCC19977) of all isolates using SeqKit (53). After subspecies identification, the maximum-likelihood phylogeny within each ABS and MAS was constructed as described above. The complete genome sequences of ATCC19977 and JCM15300 were used as a reference, respectively. Alignments containing 76,114 and 48,718 recombination-free variable positions located on their core-genome regions (3,963,788 bp for ABS and 4,033,769 bp for MAS) were used to estimate maximum-likelihood phylogenies, respectively. The resulting phylogenetic trees were used to identify clusters (ABS-EA and MAS-EA clusters) in Japan and Taiwan using the TreeGubbins software with options ‘-s (significance cut-off level) 0.01, -p (number of permutations to run to test significance) 10000’ (https://github.com/simonrharris/tree_gubbins). Only detected clusters from more than one location and greater than three isolates were considered ABS-EA/MAS-EA clusters to exclude possible point source outbreaks.

To assess the relationships among Japanese (around Tokyo and Okinawa), Taiwanese, and circulating clones of ABS or MAS in a total of eight countries, we combined our data set with publicly available WGS data from 476 MABC clinical isolates (349 ABS and 127 MAS, Table S3). These were analyzed in three previous WGS-based epidemiological studies (16, 20, 32), with one strain per patient and known cluster information to which each isolate belongs (A. Floto, J. Parkhill, and J. Bryant, personal communication). All additional raw read data were obtained from the Sequence Read Archive (SRA) and assembled *de novo* using the Shovill pipeline. To measure the assembly quality of these additional isolates, we calculated the genome fraction (%) of each isolate, which is the total number of aligned bases in the reference genomes (ATCC19977 or JCM15300) using QUAST software (54). We used only isolates with genome fractions greater than 85% for downstream analyses (Fig. S4). Alignments containing 102,613 and 58,975 recombination-free variable positions located on their core-genome regions (3,584,621 bp for ABS and 3,789,583 bp for MAS) were used to estimate maximum-likelihood phylogenies, and cluster analysis was performed as described above.

### Analysis of lineage-associated genes

Lineage-associated genes were identified as described previously (15). In brief, we annotated all draft genomes and complete genome sequences of the three type strains (ATCC19977, JCM15300, and BD) of MABC using DFAST_core ver. 1.0.3 with default settings (55) and Roary software with default settings (56) to compute core genes or accessory genes. We identified accessory genes that were significantly associated with lineage using the Scoary (57).

### Statistics

Statistical analyses were performed with R software (www.r-project.org). The R function multinom() was used to statistically assess the differences in the composition of MABC subspecies at the three locations. The R function wilcox.exact() was used to assess the statistical significance of the differences in nucleotide diversities and pairwise SNP distances among MABC clinical isolates. Enrichments of macrolide resistance-associated mutations in ABS/MAS-EA clusters were statistically assessed using the R function fisher.test().

### Study approval

This study was reviewed and approved by the Medical Research Ethics Committee of the Fukujuji Hospital (#18038), Keio University Hospital (#20080131), Okinawa Chubu Hospital (#2018-89), and National Taiwan University Hospital (#201808087RINA) for the use of human subjects.

## Data Availability

All data produced in the present study are available upon reasonable request to the corresponding authors.

## Author contributions

SM, YH, MA, JYC and PRH contributed to the design of this study; JYC, PRH, RJ, KM, KF, TK, HN, NH, MA, and SM collected the isolates used in this study; Y Mura, Y Mori, AA, and SM obtained WGS data of newly sequenced isolates; MY and JYC analyzed and interpreted data, produced figures and tables; MY and YH drafted the manuscript, and all the other authors critically revised it.

## Acknowledgments

This work was supported in part by grants from the Japan Agency for Medical Research and Development (AMED) to SM (18fk0108043h0402 and 19fk0108043h0403), MA (20fk0108129 and 21fk0108139j0902), and YH (jp20fk0108043, jp20fk0108064, jp20fk0108075, jp20fk0108093, jp21fk0108129, jp21fk0108608, jp20jm0510004, jp20wm0125007, jp20wm0225004 and jp20wm0325003), grants-in-aid from the Japan Society for Fostering Joint International Research (B) to YH and MY (jp19KK0217), for Early-Career Scientists to MY (jp20K17205), and grants from Taiwan Ministry of Science and Technology for PRH (107-2314-B-002-211 and 108-2320-B-002-047) and JYC (107-2314-B-002-245). The funders had no role in the study design, data collection and analysis, decision to publish, or preparation of the manuscript.

The clinical isolates used in this study were obtained from the hospitals and universities listed below. We appreciate the work of all clinicians at the following institutions who cared for patients infected with these mycobacteria: Fukujuji Hospital Japan Anti-Tuberculosis Association (JATA), Keio University Hospital, National Taiwan University Hospital, and Okinawa Chubu Hospital. The experiments were performed in part on the NIG Supercomputer at the ROIS National Institute of Genetics. We appreciate Tetsuya Hayashi and Yoshitoshi Ogura for technical supports on the phylogenetic analysis of MABC. We also thank Andres Floto, Julian Parkhill, and Josephine Bryant for discussions on the phylogenetic analysis of MABC.

