## Supplementary material for "Molecular epidemiological characteristics of *Mycobacterium abscessus* complex in non-cystic fibrosis patients in Japan and Taiwan": Figures S1-S4

**Supplemental Figures**

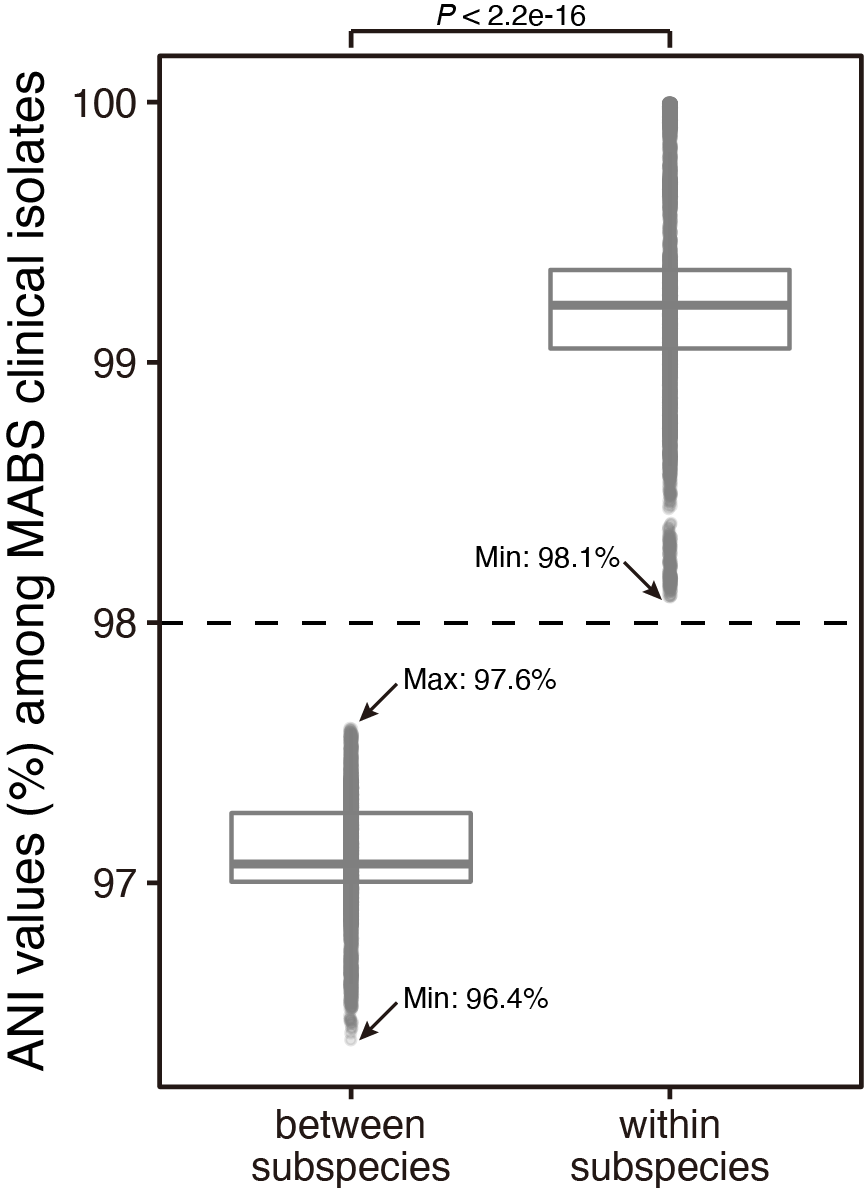

**Figure S1.** Average nucleotide identity (ANI) values among MABC clinical isolates from Japan and Taiwan. ANI values among 220 clinical isolates and three reference strains (ATCC19977, JCM15300, and BD) were measured for all strain pairs using fastANI (37). A boxplot indicates the distribution of ANI values within or between MABC subspecies. The Mann–Whitney U-test was performed to assess statistical significance between ANI values within or between the subspecies.

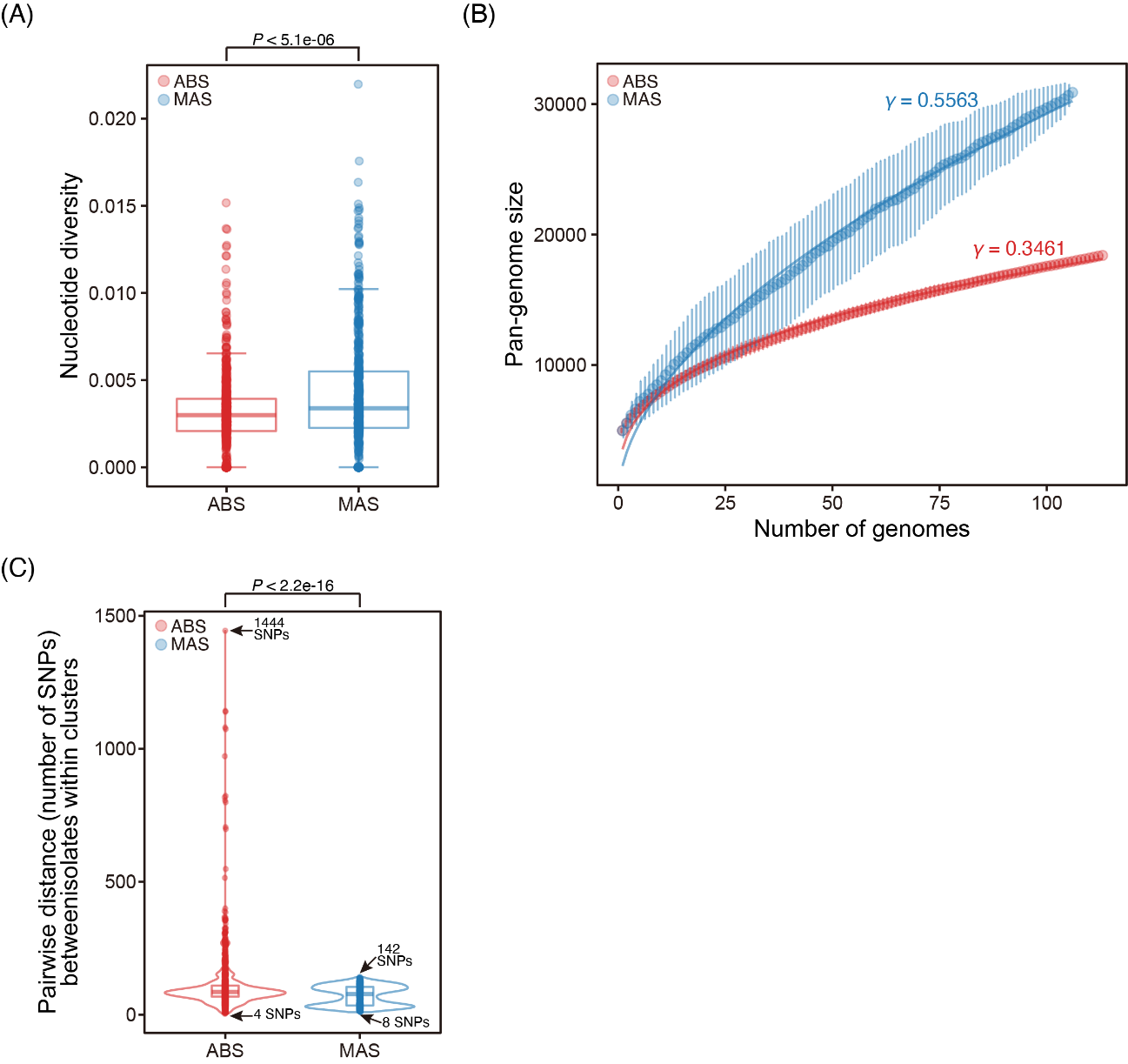

**Figure S2.** Populational comparison between ABS and MAS in Japan and Taiwan. (A) Nucleotide diversity of ABS or MAS clinical isolates in Japan and Taiwan was calculated for 10 kb nonoverlapping sliding windows of the genome alignments using the R package PopGenome (57). The Mann–Whitney U-test was used to assess the statistical significance of the differences between nucleotide diversities. (B) Comparison of pan-genome size between ABS and MAS clinical isolates in Japan and Taiwan. Orthologous gene families were identified using Roary software (41). γ denotes the parameter estimate obtained by data fitting to a power model *n* = κ×N^γ, where *n* is the number of gene families, N is the number of sampled genomes, and κ and γ are coefficients. (C) Pairwise SNP distances among clinical isolates belonging to each of six ABS-EA or five MAS-EA clusters. Whole-genome alignments containing recombination-free variable positions located in core genomes were used to calculate the SNP distances using snp-dist (https://github.com/tseemann/snp-dists). The Mann–Whitney U-test was used to assess the statistical significance of the differences in pairwise SNP distances between clinical isolates comprising the ABS- and MAS-EA clusters.

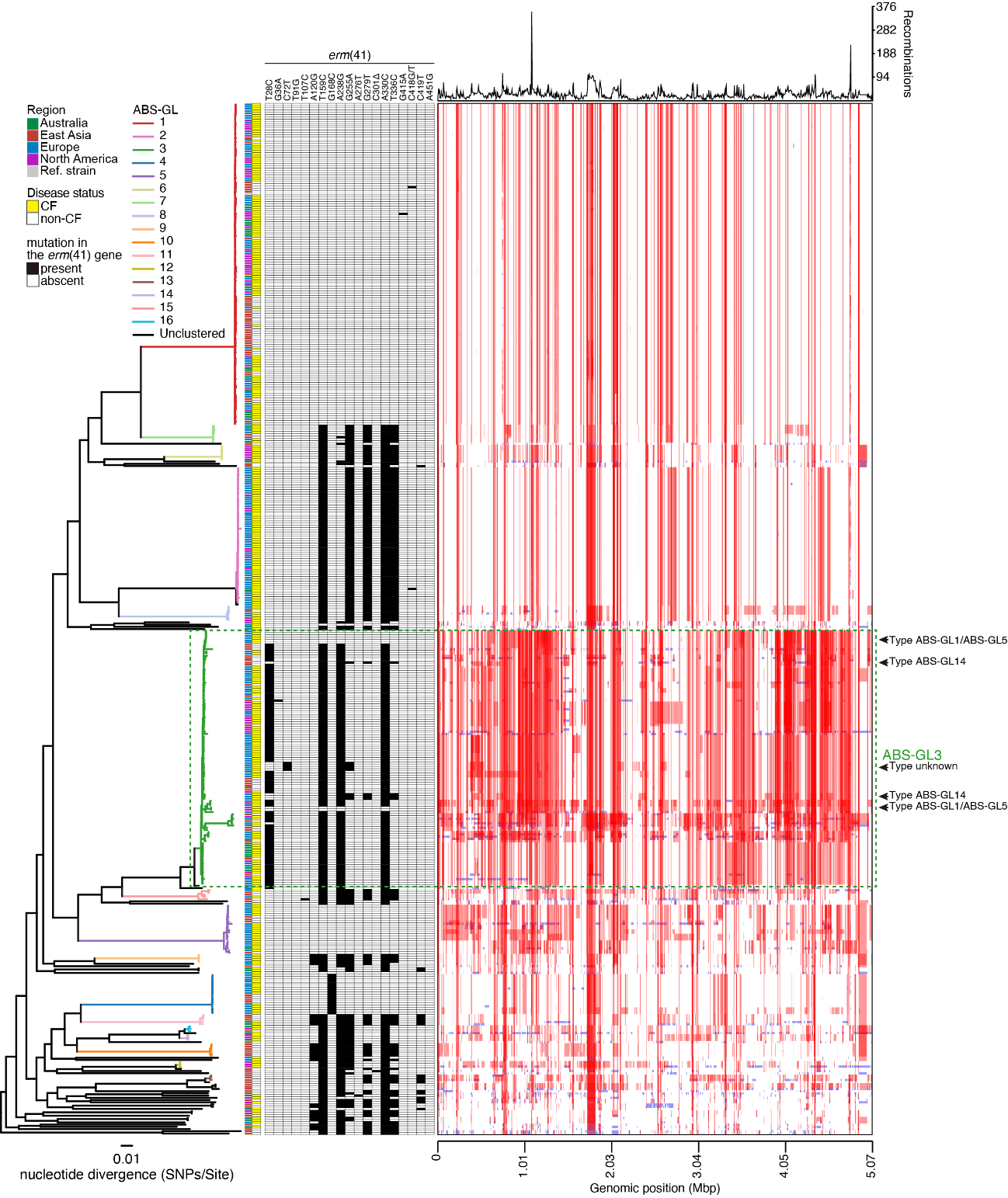

**Figure S3.** *erm*(41) gene among ABS clinical isolates. Overview of recombination events in a total of 461 ABS clinical isolates. Recombination events in internal branches (red boxes) were present in multiple clinical isolates and were shared through clonal descent, while those in the terminal branches (blue boxes) were clinical isolate-specific and represent independent recent acquisitions. Phylogenetic trees were estimated as described in Fig. 4. The presence and absence of mutations in the *erm*(41) gene, disease status (CF or non-CF) of corresponding patients, and the region where the clinical isolate was obtained are shown as Fig. 4.

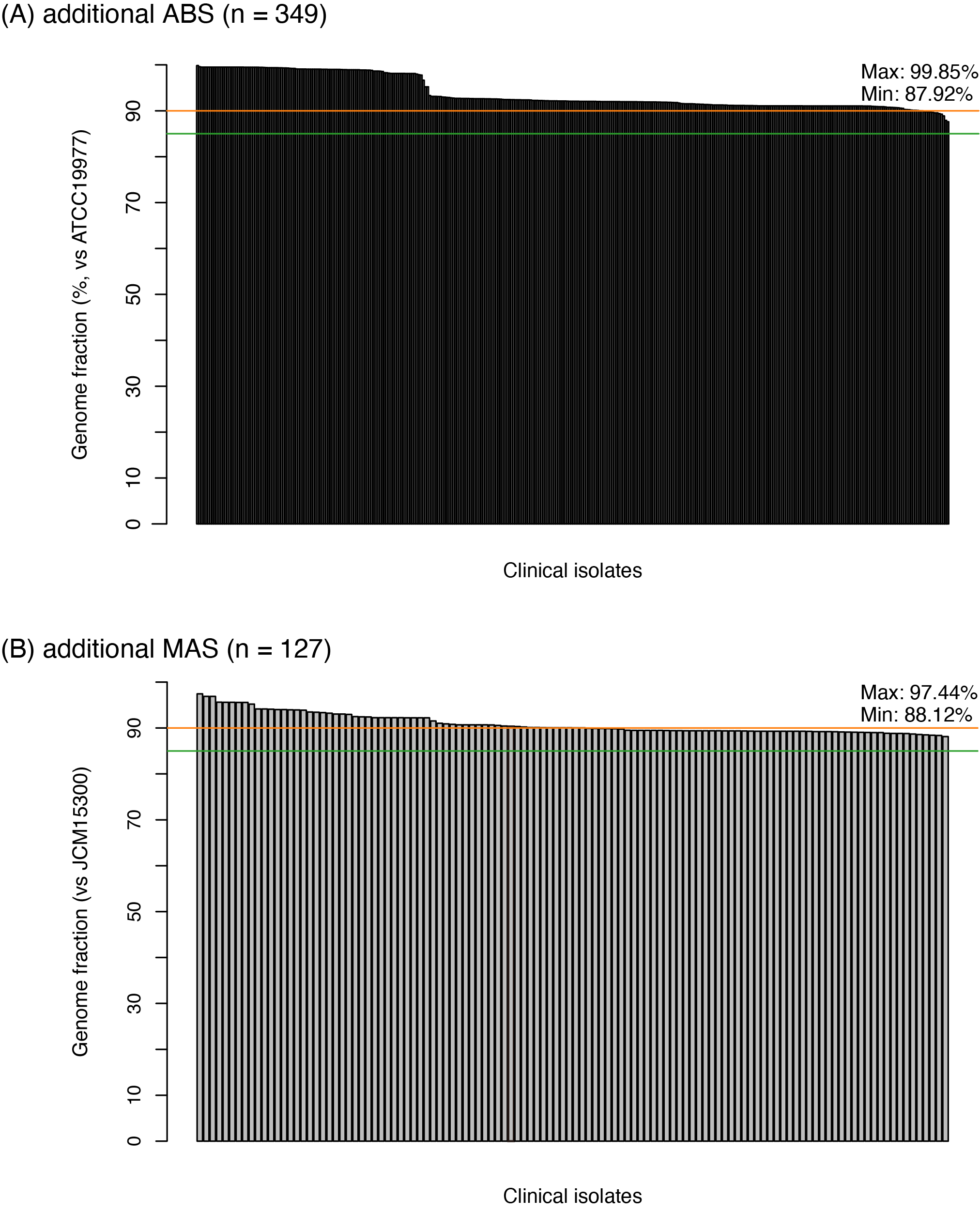

**Figure S4.** Assemble qualities of additional ABS/MAS clinical isolates. Genome fraction (%) of each additional isolate, 349 ABS (A) or 127 MAS (B), which indicates the total number of aligned bases against ATCC19977 and JCM15300, are shown, respectively. Orange and green lines indicate 90% and 85% genome fractions, respectively.
